# Impact of SGLT2 Inhibitors on Survival in Gastrointestinal Cancer Patients Undergoing Chemotherapy and/or Radiotherapy: A Real-World Data Retrospective Cohort Study

**DOI:** 10.1101/2025.01.10.25320355

**Authors:** Lucas E. Flausino, Alexis Germán Murillo Carrasco, Tatiane Katsue Furuya, Wen-Jan Tuan, Roger Chammas

## Abstract

**Background:** The role of sodium-glucose co-transporter 2 inhibitor (SGLT2i) drugs in the management of diabetes and cardiovascular disease is well-established, but emerging evidence suggests potential effects on cancer outcomes, including gastrointestinal (GI) cancers. We conducted an extensive, sex-oriented, real-world data analysis to investigate whether SGLT2i can enhance GI cancer outcomes when used alongside standard therapies such as chemotherapy and radiotherapy.

**Methods:** The study applied a retrospective cohort design with data from the TriNetX research database (https://trinetx.com), examining GI cancer patients treated with chemotherapy and/or radiotherapy between 2013 and 2023. The intervention cohort consisted of Gl cancer patients who received SGLT2i, while the control cohort did not. A 5-year follow-up period was used, and baseline characteristics were balanced using a 1:1 propensity score matching technique. Cox proportional-hazards and logistic regression models assessed mortality and morbidity risks between the cohorts.

**Results:** The study included 6,389 male and 3,457 female patients with GI cancer (ICD-10: C15-C25). The use of SGLT2i was significantly associated with improved survival for both male (HR 0.568; 95% CI 0.534-0.605) and female (HR 0.561; 95% CI 0.513-0.614) patients undergoing chemotherapy and/or radiotherapy. SGLT2i use also correlated significantly with lower hospitalisation rates both in male (OR 0.684; 95% CI 0.637-0.734) and female (OR, 0.590; 95% CI 0.536-0.650) patients. The analysis of GI cancer subtypes also demonstrated similar benefits, without significant adverse effects.

**Conclusions:** Repurposing SGLT2 inhibitors for cancer treatment could potentially improve outcomes for GI cancer patients without causing significant side effects. Further clinical trials are needed to confirm these findings and establish the optimal condition for its application in GI cancer treatment.

## 1. Background

Gastrointestinal (GI) tract cancers represent a significant proportion of the global cancer burden, accounting for over 25% of cancer diagnoses and more than 35% of cancer-related deaths (1). Recent trends indicate a notable increase in the incidence of GI cancers among younger patients, who often have a poor prognosis due to late-stage diagnoses. Investigations into various GI cancers, including colorectal, gastric, pancreatic, liver, and biliary tract cancers, have linked the early onset of these conditions to modern lifestyle factors such as obesity, high glycemic load diets, and sedentary behavior (2). In addition to these environmental factors, intrinsic mechanisms contribute to GI tumorigenesis, including adaptations in the tumor microenvironment, alterations in the immunological profile, and reprogramming of metabolic pathways to support tumor growth and invasion (3).

In this context, tumor metabolism is vital in the onset of GI cancer, highlighting it as a valuable target for potential therapies. This energetic rewire is an essential cancer hallmark for tumor cells thriving (4). Based on the Warburg effect, wherein cancer cells preferentially utilize glycolysis for energy production despite the presence of oxygen, this metabolic shift is responsible for supplying energy and metabolic intermediates needed for biosynthesis, facilitating unrestrained cell growth (5).

Multiple mechanisms contribute to this reprogramming. For instance, the activation of the Ras-PI3K-AKT-mTOR pathway results in elevated levels of glucose transporters (GLUTs), especially GLUT1, and glycolytic enzymes, which have been correlated with poor prognosis in diverse GI cancers, such as esophagus, gastric, colorectal, pancreatic, liver, and gallbladder cancers (5,6). Ultimately, these metabolic adaptations increase hypoxia, and levels of acidity, resulting in genetic and epigenetic changes that give rise to diverse cancer cell phenotypes, promoting tumor aggressiveness, immune evasion, and contributing to resistance against chemoradiotherapy (7).

Therefore, preventing glucose from entering the cell is an attractive strategy to overcome treatment resistance. In fact, glucose transporters are overexpressed in most GI cancer cells, and exploring specific inhibitors that can block cancer cells’ glucose uptake could be beneficial (6). This approach, however, has been facing challenges. For example, inhibiting GLUT1, though an appealing option to treat cancer, could potentially be problematic because normal cells also express this transporter, and its inhibition would likely lead to various detrimental side effects on healthy cells and tissues (8).

In light of that, targeting more selective glucose transporters in cancer cells might be a better option to slow tumor growth safely. Several GI cancer cells, such as those of liver, pancreatic, and colon cancer, have recently been reported to overexpress sodium-glucose co-transporter 2 (SGLT2) (9), which is a sodium-D-glucose cotransporter that functions with a 1:1 ratio of sodium to glucose. This cotransporter is highly expressed in the kidneys, where it is responsible for over 90% of the filtered D-glucose reabsorption in the proximal tubules (10), and it was shown to also contribute to glucose uptake into malignant cells (11). As an example, pancreatic cells can accumulate an SGLT-specific radioactive glucose analog, and the use of SGLT2 inhibitors has been shown to block this glucose uptake, leading to reduced tumor growth in preclinical models (12). Additional research suggests that SGLT2i can impact several GI cancers, including liver, pancreatic, and colon cancers, by inhibiting tumor growth, suppressing glycolysis, and enhancing chemotherapy efficacy (9).

Given that SGLT2 expression is relatively limited compared to the widely expressed GLUT1, repurposing existing FDA-approved SGLT2 inhibitors (SGLT2i) may offer a promising avenue for contributing to cancer treatment with minimal side effects. SGLT2i were developed as antidiabetic drugs, lowering glucose plasma levels through inhibition of kidney glucose reabsorption and promoting glycosuria (13). Initially developed to treat type 2 diabetes patients, these drugs have also been shown to contribute in renal protection, weight loss, and blood pressure lowering, as well as reducing cardiovascular morbidity and mortality (14–16). Moreover, due to their relatively safe profile (17–22) and benefits in reducing cardiovascular events, they are also first-line therapy in patients with heart failure with either reduced or preserved ejection fraction, independent of their diabetes status (23,24).

Therefore, considering the established safety and benefits of SGLT2i, and their potential to interfere with GI tumor cell glycolytic metabolism, we hypothesized that these drugs could enhance patient outcomes when used alongside standard cancer therapies, including chemotherapy and/or radiotherapy. To investigate this hypothesis, we conducted an extensive analysis of real-world data from patients with various GI cancers. Utilizing the TriNetx Global Collaborative Network, a database of electronic health records (EHR), we examined the association between SGLT2i use and mortality among GI cancer patients receiving chemotherapy and/or radiotherapy. Our analysis encompassed patients with cancers of the esophagus, stomach, small intestine, colon, rectosigmoid junction, rectum, anus, liver, intrahepatic bile ducts, gallbladder, biliary tract, and pancreas. Additionally, we investigated the impact of SGLT2i on hospitalization rates and the incidence of potential adverse events.

## 2. Methods

### 2.1. Study design and data source

This study was a retrospective cohort analysis using EHR from 133 healthcare organizations (HCO) within the TriNetX Global Collaborative Network, which includes de-identified EHR data (such as demographics, diagnoses, treatments, medications, and lab results) from over 157 million patients. Data queries were conducted via the TriNetX online portal (https://trinetx.com), and the results presented only aggregated counts and statistical summaries. Since no identifiable patient data was used or accessed, the study was deemed exempt from Institutional Review Board review.

The study population consisted of individuals on radiotherapy and/or chemotherapy treated for the most prevalent GI cancers (e.g., esophagus, stomach, small intestine, colon, rectosigmoid junction, rectum, anus, liver and intrahepatic bile ducts, gallbladder, biliary tract, and pancreas) (Table S1) between December 1, 2013, and December 31, 2023. Patients were separated into two cohorts based on the use of SGLT2i, which were identified using normalized names and code sets for medications based on the Anatomical Therapeutic Chemical (ATC) system (Table S2). To account for potential biological sex differences in cancer outcomes (25–27), we analyzed cohorts consisting of either female or male patients separately.

The index event for this analysis was defined as a diagnosis of GI cancer and the initiation of cancer treatment with chemotherapy and/or radiotherapy in the control cohort. In the intervention cohort, the index event included the same criteria, with the addition of concurrent SGLT2i use. To increase the specificity of our findings, we analyzed individual queries for each GI cancer type included in the analysis, following the same eligibility and index criteria. The follow-up period began one day after the first occurrence of the index event and continued for up to five years (1825 days).

### 2.2. Outcome measures

The primary outcome measure was the 5-year mortality after the initial medication (chemotherapy and/or radiotherapy plus SGLT2i). Secondary outcomes focused on the presence of potential associated effects and complications of SGLT2i use (28), including hospitalizations, hypoglycemia, diabetic ketoacidosis, urinary tract infection, cardiovascular and cerebrovascular events, hepatic failure, acute kidney failure, and immune-related adverse events within 5 years after the medications were administered (Table S3).

### 2.3. Statistical analyses

Baseline characteristics, including patient characteristics (age, race/ethnicity), comorbidities (diabetes, ischemic heart diseases, heart failure, arterial and arteriolar diseases, cerebrovascular diseases, and chronic kidney disease), medications (metformin, insulin, antilipemic agents, and antihypertensive drugs), cancer stages, and clinical-relevant features, such as body-mass index (BMI), Hemoglobin A1C (HbA1c), NT-proBNP, Left Ventricular Ejection Fraction (LVEF), and Eastern Cooperative Oncology Group (ECOG) performance status values, were accounted for as confounding variables (see Table S4 for the full list of covariates).

The study used a 1:1 propensity score matching (PSM) approach to equalize baseline characteristics by creating matched pairs of patients with similar propensity scores from the two study groups. The PSM process was carried out using logistic regression and nearest neighbor algorithms, with a caliper width set at 0.1 times the pooled standard deviation (SD), ensuring that the matched pairs had comparable baseline characteristics.

Cox proportional hazard models were applied to assess the risk of all-cause mortality in cancer patients prescribed SGLT2i within 5 years of the initial prescription, compared to patients in the non-SGLT2i (control) cohort. Hazard ratios (HR) with 95% confidence intervals (95% CI) for the likelihood of all-cause mortality were calculated with a two-sided *p*<0.05 for statistical significance.

A logistic regression model was applied to calculate Odds ratios (OR) with 95% CI to measure the association between SGLT2i use and possible side effects and complications, with a two-sided *p*<0.05 for statistical significance. All data queries and statistical analyses were performed on the TriNetX portal. Detailed diagnosis and laboratory codes for baseline characteristics and outcome measures are available in the supplemental.

## 3. Results

### 3.1 SGLT2i use is associated with overall survival in GI male and female cancer patients

We identified 6,389 male and 3,457 female GI cancer patients who used SGLT2i under GI cancer treatment with chemotherapy and/or radiotherapy and their matched controls that did not use SGLT2i. Baseline characteristics of the study and matched control populations are shown in Table 1. In male GI cancer patients, SGLT2i use was significantly associated with a 54.99% survival rate when compared with a 35.43% survival rate of their matched control cohort at the end of the 5-year time window (HR 0.568; 95% CI 0.534-0.605). In female GI cancer patients, SGLT2i use was significantly associated with a 5-year overall survival rate of 60.63% while the matched control cohort had a 42.10% survival rate in the same follow-up period (HR 0.561; 95% CI 0.513-0.614) (Figure 1).

**Figure 1.**
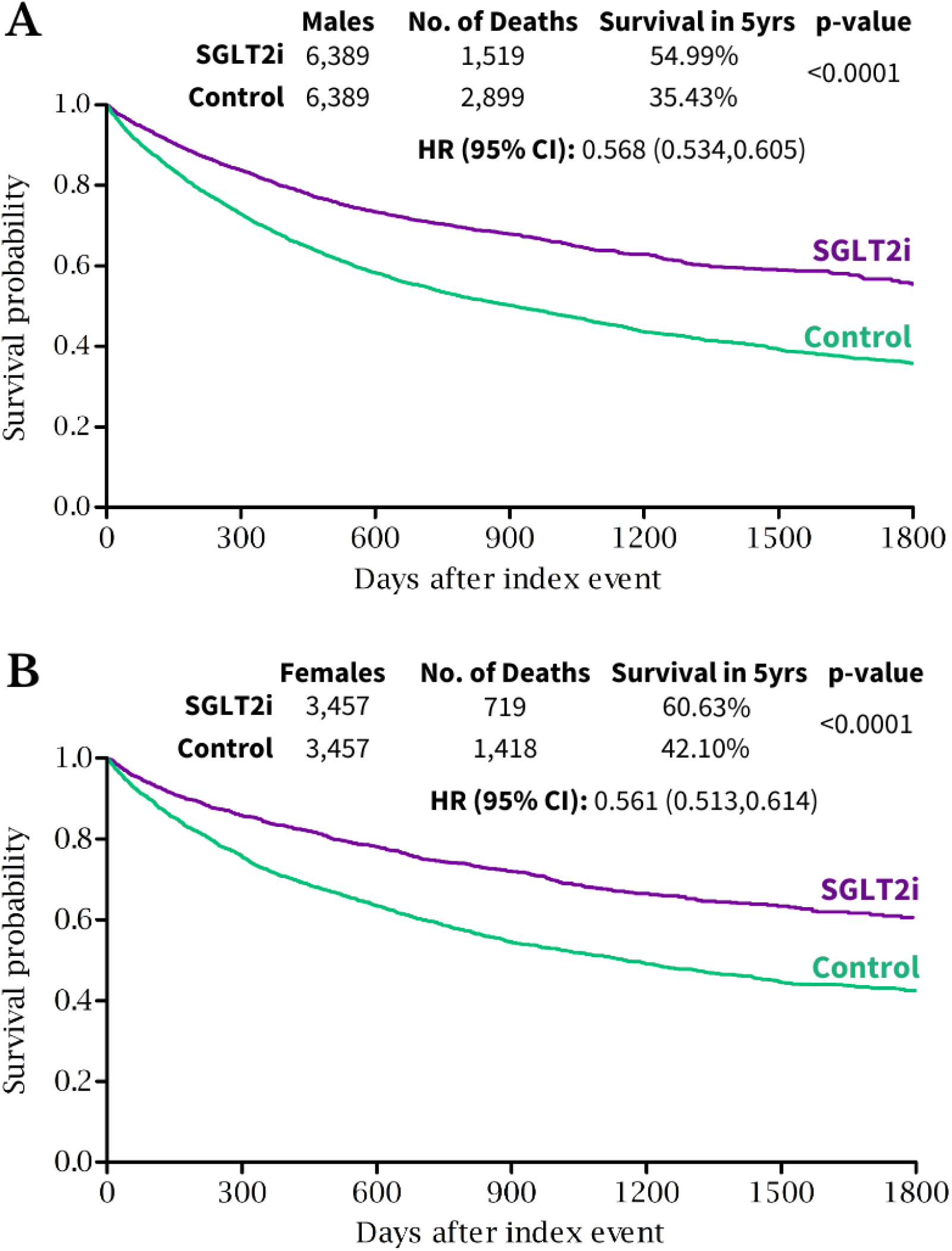
Overall Survival for gastrointestinal (GI) cancer patients that received SGLT2i under chemotherapy and/or radiotherapy regimens. **(A)** Kaplan-Meier curves of male and **(B)** female patients that used SGLT2i under GI cancer treatment. HR: Hazard ratio; CI: confidence interval; yrs: years.

**Table 1.** Propensity score-matched baseline characteristics for male and female gastrointestinal (GI) patients treated with radiotherapy and/or chemotherapy.

| Propensity Score-Matched Baseline Characteristics for Patients Treated with Radiotherapy and/or Chemotherapy |  |  |  |  |  |  |
| --- | --- | --- | --- | --- | --- | --- |
|  | Male Patients |  |  | Female Patients |  |  |
|  | SGLT2i (N=6,389) | Control (N=6,389) | P-value | SGLT2i (N=3,457) | Control (N=3,457) | P-value |
| Age at Index, mean +/- SD | 66.90 +/- 11.28 | 67.34 +/- 11.77 | 0.033 | 65.66 +/- 13.48 | 66.16 +/- 14.42 | 0.136 |
| Race and ethnicity, N (%) |  |  |  |  |  |  |
| White | 3979 (62.28%) | 4080 (63.86%) | 0.064 | 1937 (56.03%) | 2001 (57.88%) | 0.120 |
| Asian | 672 (10.52%) | 620 (9.70%) | 0.127 | 354 (10.24%) | 338 (9.78%) | 0.521 |
| Black or African American | 596 (9.33%) | 587 (9.19%) | 0.784 | 616 (17.82%) | 590 (17.07%) | 0.410 |
| Hispanic or Latino | 471 (7.37%) | 458 (7.17%) | 0.658 | 309 (8.94%) | 320 (9.26%) | 0.645 |
| Diagnosis, N (%) |  |  |  |  |  |  |
| Hypertensive diseases | 5242 (82.05%) | 5341 (83.60%) | 0.020 | 2866 (82.90%) | 2962 (85.68%) | 0.002 |
| Diabetes mellitus | 5119 (80.12%) | 5344 (83.64%) | <0.001 | 2778 (80.36%) | 2938 (84.99%) | 0.000 |
| Ischemic heart diseases | 3184 (49.84%) | 3163 (49.51%) | 0.710 | 1377 (39.83%) | 1350 (39.05%) | 0.506 |
| Diseases of arteries, arterioles and capillaries | 2055 (32.17%) | 2016 (31.55%) | 0.459 | 1104 (31.94%) | 1058 (30.61%) | 0.233 |
| Heart failure | 2004 (31.37%) | 1838 (28.77%) | 0.001 | 1088 (31.47%) | 995 (28.78%) | 0.015 |
| Chronic kidney disease (CKD) | 1995 (31.23%) | 1901 (29.75%) | 0.071 | 1022 (29.56%) | 999 (28.90%) | 0.543 |
| Cerebrovascular diseases | 1149 (17.98%) | 1147 (17.95%) | 0.963 | 701 (20.28%) | 667 (19.29%) | 0.305 |
| Laboratory values, mean +/- SD |  |  |  |  |  |  |
| BMI | 28.89 +/- 6.36 | 28.40 +/- 6.24 | < 0.001 | 30.16 +/- 7.79 | 29.28 +/- 7.76 | <0.001 |
| HbA1c (%) | 7.37 +/- 1.79 | 6.93 +/- 1.70 | < 0.001 | 7.44 +/- 1.80 | 6.92 +/- 1.76 | <0.001 |
| NT-proBNP [Mass/volume] | 2915.91 +/- 5912.30 | 2536.26 +/- 6149.77 | 0.183 | 2733.47 +/- 5387.39 | 2405.17 +/- 5837.12 | 0.344 |
| LVEF (%) | 51.55 +/- 15.06 | 55.26 +/- 13.48 | < 0.001 | 55.02 +/- 14.12 | 60.17 +/- 12.45 | <0.001 |
| ECOG Performance Status | - | - | - | - | - | - |
| Medications, N (%) |  |  |  |  |  |  |
| Antilipemic agents | 4905 (76.77%) | 4955 (77.56%) | 0.292 | 2563 (74.14%) | 2585 (74.78%) | 0.544 |
| Beta blockers | 4477 (70.07%) | 4493 (70.32%) | 0.757 | 2295 (66.39%) | 2241 (64.83%) | 0.172 |
| Insulin | 4309 (67.44%) | 4425 (69.26%) | 0.027 | 2374 (68.67%) | 2410 (69.71%) | 0.348 |
| Diuretics | 4107 (64.28%) | 4091 (64.03%) | 0.768 | 2307 (66.73%) | 2314 (66.94%) | 0.858 |
| Metformin | 3768 (58.98%) | 3712 (58.10%) | 0.315 | 2024 (58.55%) | 1945 (56.26%) | 0.055 |
| Calcium channel blockers | 3193 (49.98%) | 3153 (49.35%) | 0.479 | 1740 (50.33%) | 1741 (50.36%) | 0.981 |
| ACE inhibitors | 2989 (46.78%) | 3063 (47.94%) | 0.190 | 1539 (44.52%) | 1547 (44.75%) | 0.847 |
| Angiotensin II inhibitors | 2705 (42.34%) | 2600 (40.70%) | 0.059 | 1495 (43.25%) | 1411 (40.82%) | 0.041 |
| Oncology, N (%) |  |  |  |  |  |  |
| Stage 0 | 30 (0.47%) | 22 (0.34%) | 0.266 | 19 (0.55%) | 18 (0.52%) | 0.869 |
| **Stage 1** | 263 (4.12%) | 254 (3.98%) | 0.686 | 212 (6.13%) | 210 (6.08%) | 0.920 |
| **Stage 2** | 315 (4.93%) | 309 (4.84%) | 0.805 | 161 (4.66%) | 159 (4.60%) | 0.909 |
| **Stage 3** | 330 (5.17%) | 297 (4.65%) | 0.177 | 155 (4.48%) | 138 (3.99%) | 0.310 |
| **Stage 4** | 230 (3.60%) | 237 (3.71%) | 0.741 | 109 (3.15%) | 98 (2.84%) | 0.438 |

N: number of individuals; SD: standard deviation; HbA1c: Hemoglobin A1c in the blood; NT-proBNP: N-terminal pro-B-type natriuretic peptide in Serum, Plasma or Blood; LVEF: Left Ventricular Ejection Fraction; BMI: body-mass index; ACE: angiotensin-converting enzyme.

### 3.2 SGLT2i use association with secondary outcomes in male and female GI cancer patients

SGLT2i use in conjunction with cancer treatment was significantly associated with a reduction in the rates of hospitalization, hypoglycemic events, urinary tract infections, acute kidney failure, and hepatic failure. Moreover, male patients in the SGLT2i cohort were associated with fewer immune-related adverse events. In contrast, there was no significant association between SGLT2i and cardiovascular and cerebrovascular events and immune-related adverse events in female patients. Additionally, ketoacidosis events were more significantly frequent in male patients who used SGLT2i while on cancer treatment, whereas there was no significant association with these events in female patients (Figure 2).

**Figure 2.**
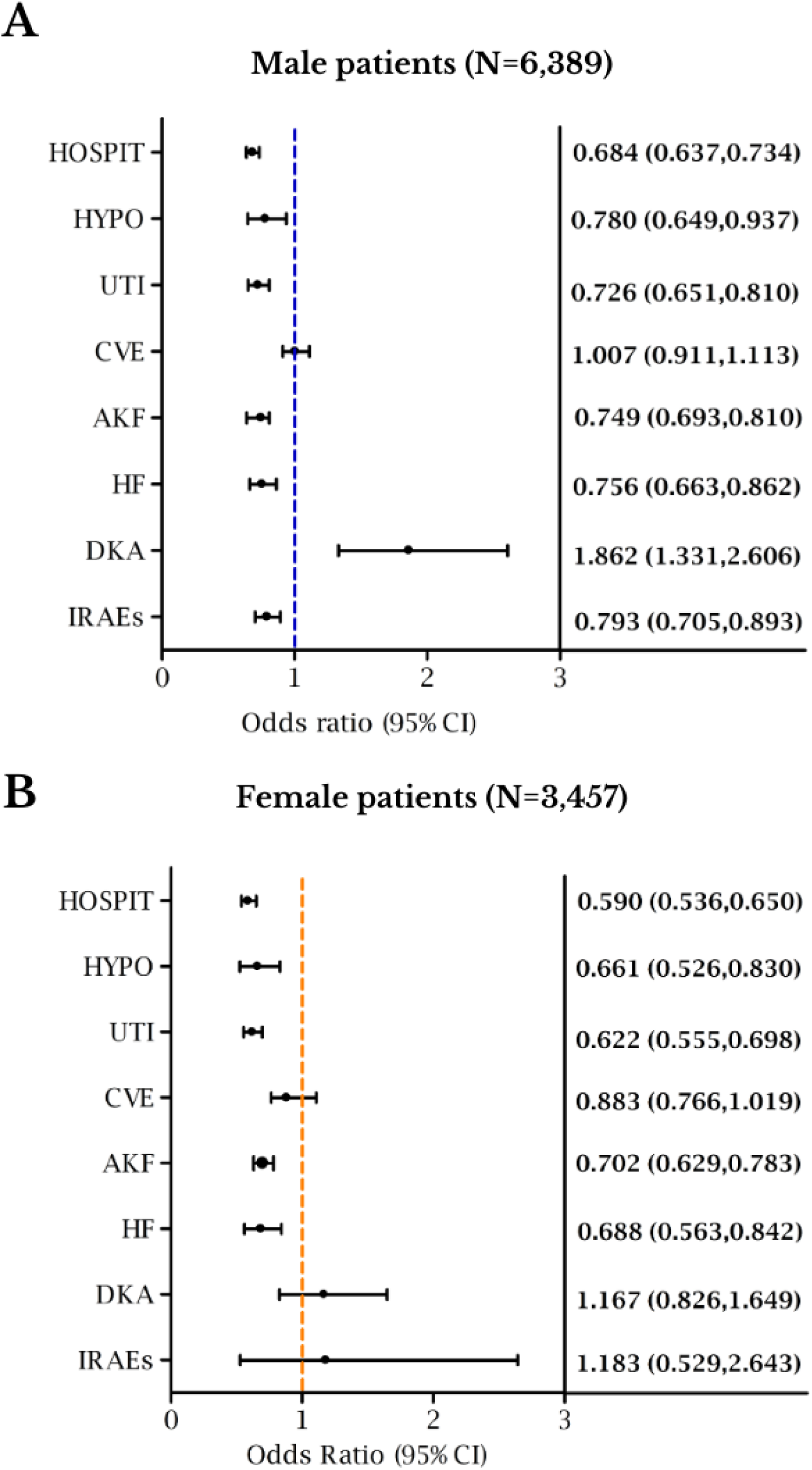
Association of SGLT2i use with gastrointestinal (GI) cancer patients secondary outcomes. HOSPIT, hospitalisation; HYPO, hypoglycemic events; UTI, urinary tract infections; CVE, cardiovascular and cerebrovascular events; AKF, acute kidney failure; HF, hepatic failure; DKA, diabetic ketoacidosis events; IRAEs, immune-related adverse events; CI: confidence interval; N: number of individuals.

### 3.3 SGLT2i use during GI cancer treatment is associated with specific GI cancer types reduced mortality and hospitalization

The use of SGLT2i with chemotherapy and/or radiotherapy correlated significantly with increased survival in male patients with esophagus, stomach, colon, rectosigmoid junction, rectum, anus and canal anal, liver and intrahepatic bile duct, and pancreatic cancers. On the other hand, there was no significant association between SGLT2i and survival of the small intestine, gallbladder, and biliary tract in male cancer patients (Figure 3A).

**Figure 3.**
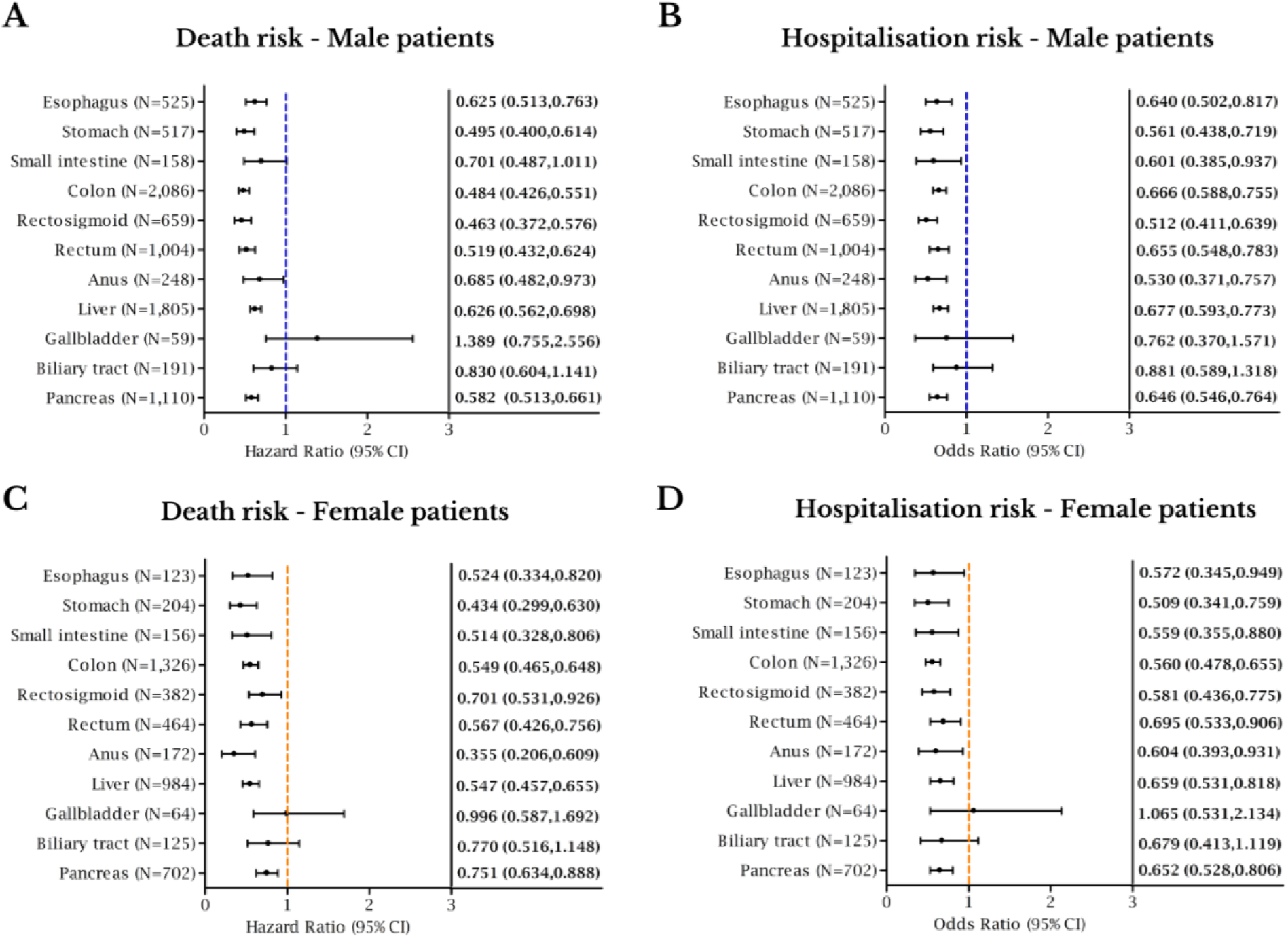
The association between the use of SGLT2i with death and hospitalisation risks in different types of gastrointestinal cancer. CI: confidence interval; N: number of individuals.

Moreover, hospitalization rates were diminished in male patients with esophagus, stomach, small intestine, colon, rectosigmoid junction, rectum, anus and canal anal, liver and intrahepatic bile duct, and pancreatic cancers that used SGLT2i. However, hospitalization did not correlate with SGLT2i use in the gallbladder or biliary tract in male cancer patients (Figure 3B).

In females, SGLT2i use demonstrated an increased association with reduced mortality in patients with esophagus, stomach, small intestine, colon, rectosigmoid junction, rectum, anus and canal anal, liver and intrahepatic bile duct, and pancreatic cancers. Meanwhile, gallbladder and biliary tract cancer patients did not significantly benefit from SGLT2i during their treatments (Figure 3C).

Furthermore, SGLT2i was significantly associated with fewer hospitalization events in GI female cancer patients, specifically in those with esophagus, stomach, small intestine, colon, rectosigmoid junction, rectum, anus and canal anal, liver and intrahepatic bile duct, and pancreatic cancers. In contrast, the incidence of these events in gallbladder and biliary tract cancer female patients who used SGLT2i was not significant (Figure 3D).

### 3.4 SGLT2i use alongside GI cancer treatment has a safe profile and does not associate with adverse events

The use of SGLT2i in combination with chemotherapy and/or radiotherapy was not linked to an increased risk of adverse treatment events for any of the specific GI cancer types studied. In male patients, SGLT2i were associated with reduced hypoglycemic events in colon, liver and intrahepatic bile duct cancer patients. In addition, its use correlated with reduced urinary tract infections in the small intestine, colon, rectum, anus and canal anal, and liver and intrahepatic bile ducts cancer patients, as well as reduced acute kidney failure events in esophagus, stomach, colon, rectosigmoid junction, rectum, anus, canal anal, liver and intrahepatic bile ducts, biliary tract, and pancreatic cancer patients. Moreover, SGLT2i was protective against cardiovascular and cerebrovascular events, and hepatic failure in liver and intrahepatic bile ducts cancer patients, and reduced immune-related adverse events in patients with colon, rectosigmoid junction, rectum, anus and canal anal cancers (Table 2).

**Table 2.** Risks of secondary outcomes according to different types of gastrointestinal (GI) cancer.

| Risks of secondary outcomes according to different types of GI cancer |  |  |  |  |  |  |  |
| --- | --- | --- | --- | --- | --- | --- | --- |
| Male - Odds Ratio (95% CI) |  |  |  |  |  |  |  |
| GI cancer type | Hypoglycemia | Urinary Tract Infection | Cardiovascular and cerebrovascular events | Acute Kidney Failure | Hepatic Failure | Ketoacidosis | Immune-related adverse events |
| **Esophagus (N=525)** | 0.759<br>(0.365,1.578) | 0.898<br>(0.598,1.349) | 1.014 (0.728,1.415) | 0.704<br>(0.530,0.934) | 1.205<br>(0.516,2.813) | 1.000<br>(0.413,2.423) | 0.829<br>(0.526,1.306) |
| **Stomach (N=517)** | 0.730<br>(0.404,1.320) | 0.901<br>(0.604,1.345) | 0.714 (0.509,1.000) | 0.708<br>(0.535,0.937) | 0.676<br>(0.33,1.384) | 1.000<br>(0.413,2.423) | 1.115<br>(0.706,1.760) |
| **Small intestine (N=158)** | 1.000<br>(0.404,2.474) | 0.459<br>(0.221,0.954) | 1.151 (0.631,2.097) | 0.886<br>(0.547,1.435) | 1.000<br>(0.404,2.474) | 1.000<br>(0.404,2.474) | 0.494<br>(0.222,1.100) |
| **Colon (N=2,086)** | 0.580<br>(0.413,0.815) | 0.615<br>(0.513,0.737) | 1.036 (0.871,1.231) | 0.632<br>(0.550,0.726) | 0.791<br>(0.559,1.120) | 1.176<br>(0.672,2.058) | 0.771<br>(0.633,0.939) |
| **Rectosigmoid junction (N=659)** | 0.876<br>(0.489,1.570) | 0.766<br>(0.564,1.040) | 0.911 (0.675,1.229) | 0.594<br>(0.468,0.755) | 0.857<br>(0.496,1.479) | 1.372<br>(0.625,3.010) | 0.683<br>(0.493,0.946) |
| **Rectum (N=1,004)** | 0.829<br>(0.484,1.420) | 0.630<br>(0.498,0.798) | 1.140 (0.894,1.454) | 0.641<br>(0.527,0.781) | 0.613<br>(0.361,1.039) | 1.392<br>(0.678,2.856) | 0.672<br>(0.498,0.905) |
| **Anus and canal anal (N=248)** | 0.826<br>(0.350,1.949) | 0.599<br>(0.373,0.962) | 0.925 (0.591,1.447) | 0.514<br>(0.353,0.749) | 1.000<br>(0.409,2.447) | 1.000<br>(0.409,2.447) | 0.295<br>(0.162,0.536) |
| **Liver and intrahepatic bile ducts (N=1,805)** | 0.651<br>(0.470,0.901) | 0.734<br>(0.598,0.901) | 0.794 (0.656,0.961) | 0.684<br>(0.595,0.786) | 0.686<br>(0.583,0.809) | 1.540<br>(0.801,2.962) | 0.967<br>(0.769,1.215) |
| **Gallbladder (N=59)** | 1.000<br>(0.382,2.616) | 1.000<br>(0.382,2.616) | 1.000 (0.382,2.616) | 1.085<br>(0.492,2.391) | 1.000<br>(0.382,2.616) | NC | 1.000<br>(0.382,2.616) |
| **Other and unspecified parts of biliary tract (N=191)** | 1.000<br>(0.406,2.461) | 0.949<br>(0.503,1.790) | 1.239 (0.651,2.359) | 0.590<br>(0.379,0.919) | 0.64<br>(0.315,1.300) | 1.000<br>(0.406,2.461) | 1.062<br>(0.539,2.092) |
| **Pancreas (N=1,110)** | 1.000<br>(0.702,1.425) | 0.928<br>(0.697,1.236) | 1.201 (0.942,1.531) | 0.779<br>(0.642,0.944) | 0.928<br>(0.634,1.357) | 1.682<br>(0.882,3.208) | 0.979<br>(0.737,1.301) |
| Female - Odds Ratio (95% CI) |  |  |  |  |  |  |  |
| GI cancer type | Hypoglycemia | Urinary Tract Infection | Cardiovascular and cerebrovascular events | Acute Kidney Failure | Hepatic Failure | Ketoacidosis | Immune-related adverse events |
| **Esophagus (N=123)** | 1.000<br>(0.401,2.496) | 0.659<br>(0.348,1.247) | 0.697 (0.352,1.382) | 0.754<br>(0.431,1.318) | 1.000<br>(0.401,2.496) | 1.000<br>(0.401,2.496) | 1.422<br>(0.678,2.984) |
| **Stomach (N=204)** | 0.649<br>(0.285,1.482) | 0.898<br>(0.570,1.415) | 0.604 (0.340,1.075) | 0.802<br>(0.519,1.239) | 1.000<br>(0.407,2.457) | 1.000<br>(0.407,2.457) | 0.855<br>(0.494,1.481) |
| **Small intestine (N=156)** | 1.000<br>(0.404,2.475) | 0.673<br>(0.386,1.173) | 1.000 (0.499,2.003) | 0.739<br>(0.431,1.269) | 1.000<br>(0.404,2.475) | 1.000<br>(0.404,2.475) | 1.000<br>(0.551,1.814) |
| **Colon (N=1,326)** | 0.750<br>(0.503,1.119) | 0.524<br>(0.437,0.628) | 0.754 (0.605,0.940) | 0.573<br>(0.480,0.686) | 1.032<br>(0.631,1.689) | 1.194<br>(0.665,2.144) | 0.832<br>(0.670,1.034) |
| **Rectosigmoid junction (N=382)** | 0.756<br>(0.362,1.580) | 0.663<br>(0.481,0.914) | 0.852 (0.575,1.262) | 0.533<br>(0.388,0.733) | 0.658<br>(0.292,1.483) | 1.000<br>(0.411,2.431) | 0.787<br>(0.522,1.186) |
| **Rectum (N=464)** | 0.556<br>(0.297,1.042) | 0.648<br>(0.484,0.866) | 1.040 (0.705,1.535) | 0.592<br>(0.439,0.798) | 0.807<br>(0.384,1.697) | 1.000<br>(0.412,2.426) | 0.619<br>(0.417,0.921) |
| **Anus and canal anal (N=172)** | 0.903<br>(0.373,2.186) | 0.497<br>(0.304,0.813) | 1.199 (0.664,2.166) | 0.669<br>(0.417,1.073) | 1.000<br>(0.405,2.467) | 1.107<br>(0.457,2.678) | 1.841<br>(0.998,3.395) |
| **Liver and intrahepatic bile ducts (N=684)** | 0.832<br>(0.489,1.417) | 0.851<br>(0.659,1.099) | 0.748 (0.534,1.046) | 0.721<br>(0.572,0.909) | 0.757<br>(0.571,1.004) | 1.718<br>(0.781,3.779) | 1.316<br>(0.942,1.839) |
| **Gallbladder (N=64)** | 1.000<br>(0.385,2.597) | 1.228<br>(0.504,2.991) | 1.000 (0.385,2.597) | 0.915<br>(0.4,2.094) | 1.000<br>(0.385,2.597) | 1.000<br>(0.385,2.597) | 1.000<br>(0.385,2.597) |
| **Other and unspecified parts of biliary tract (N=125)** | 1.000<br>(0.401,2.494) | 1.302<br>(0.727,2.332) | 1.284 (0.575,2.867) | 0.826<br>(0.481,1.421) | 1.000<br>(0.417,2.399) | 1.000<br>(0.417,2.399) | 1.000<br>(0.401,2.494) |
| **Pancreas (N=702)** | 1.140<br>(0.727,1.787) | 0.606<br>(0.463,0.794) | 0.913 (0.649,1.284) | 0.687<br>(0.536,0.881) | 0.740<br>(0.444,1.233) | 1.634<br>(0.812,3.29) | 0.929<br>(0.664,1.300) |

The risk of ketoacidosis in male patients with gallbladder cancer was not calculated (NC) due to the absence of events in the cohorts. N: number of individuals; CI: confidence interval; GI: gastrointestinal.

In female patients, SGLT2i were associated with reduced urinary tract infection in colon, rectosigmoid junction, rectum, anus and canal anal, and pancreatic cancers patients, as well as fewer cardiovascular and cerebrovascular events in colon cancer patients. Furthermore, patients with colon, rectosigmoid junction, rectum, liver and intrahepatic bile ducts, and pancreatic cancers, presented fewer acute kidney injuries when compared with their respective control cohorts. Additionally, rectal cancer patients who used SGLT2i, while on cancer treatment, experienced significantly fewer immune adverse events than their controls (Table 2).

## 4. Discussion

Our findings indicate that the use of SGLT2i is significantly associated with improved GI cancer outcomes. Specifically, SGLT2i use appears to reduce overall mortality, hospitalization rates, and adverse events both in male and female GI cancer patients. This effect may be mediated through the inhibition of glucose uptake by tumor cells, thereby altering their metabolism and survival. As a result, repurposing SGLT2i may enhance the effectiveness of chemotherapy and/or radiotherapy in GI cancer treatment.

Besides reducing systemic glucose levels and controlling glucose uptake by cancer cells, SGLT2i can potentially interfere with multiple signaling pathways. In liver cancer, preclinical studies demonstrated that the use of canagliflozin, an SGLT2i, promoted proteasomal degradation, downregulated glycolytic and fatty acid metabolism, attenuated angiogenic activity, targeted the AMPK/mTOR pathway, and regulated the endoplasmic reticulum (ER) stress-mediated autophagy, inhibiting the proliferation and invasion of hepatocellular carcinoma cells (29–33). Moreover, canagliflozin has cancer treatment adjuvant properties, sensitizing tumor liver cells to radiotherapy (33) and chemotherapy (34,35) as well as to specific treatments such as sorafenib (36).

In preclinical models of pancreatic cancer, canagliflozin and dapagliflozin reduced cell proliferation, suppressed tumor glycolysis, induced tumor necrosis, and enhanced the effects of gemcitabine chemotherapy (12,37,38). Additionally, these inhibitors reversed hyperinsulinemia, impaired cell adhesion, and induced mitochondrial dysfunction and ER-stress autophagy, contributing to slower tumor growth in colon cancer models (39–41). SGLT2i also shows promise in gastric cancer, where they suppress tumor growth and metastasis by inducing ubiquitination of the tumorigenic protein YAP1 and through epigenetic modulation (42,43).

Our findings support these mechanisms, suggesting that SGLT2i could improve survival and reduce hospitalization risks for both male and female patients. However, further research is needed to clarify their impact on other GI cancers, particularly those affecting the esophagus, anus and canal anal, gallbladder, and biliary tract.

Diabetes Mellitus is a highly prevalent comorbidity among patients with GI cancers (44), with a notable association between the onset of diabetes and cancer diagnosis (45). Not only does diabetes serve as a significant risk factor for the development of various GI cancers, such as colorectal (46), gastric (47–49), pancreatic (50), and liver (51) cancers, but cancer patients are also at an increased risk of developing new-onset type 2 diabetes, often requiring insulin therapy to manage their condition (52,53). This complex interplay between type 2 diabetes and cancer is shaped by metabolic abnormalities such as hyperinsulinemia, elevated insulin-like growth factor I (IGF-I), hyperglycemia, and inflammatory cytokines (54). These factors not only accelerate cancer progression but also increase mortality rates in male and female patients with colon, liver, and intrahepatic bile duct, and pancreatic cancers (55).

In addition, cancer can disrupt metabolic interactions in peripheral tissues, leading to insulin resistance and redirecting glucose from skeletal muscle and adipose tissue to tumor cells, fueling their growth and invasion (56). This energetic shunt plays a key role in cancer cachexia (57,58), a debilitating and often fatal condition marked by severe muscle and fat loss, and commonly present in GI tumors, such as in colorectal, gastroesophageal, hepatobiliary and pancreatic cancers (59–61).

In this context, the use of SGLT2i may offer significant benefits in improving cancer treatment outcomes by potentially reducing the tumor’s metabolic advantage and fostering a healthier physiological environment that enhances both cancer immunosurveillance and treatment tolerance. As an example, in GI cancer patients with diabetes, SGLT2i may help achieve better glycemic control and reduce hyperinsulinemia, without major side effects, decreasing growth stimuli for cancer cells (54) while also improving tolerance to chemotherapy and radiotherapy (62,63). Given these potential benefits, further clinical trials are needed to assess the impact of reducing insulin resistance and improving diabetic control with these drugs on outcomes in GI cancer patients.

Furthermore, SGLT2i may optimize both weight management (64,65) and blood pressure (65,66), which are conditions closely related to metabolic syndrome, a condition that usually influences GI cancer risk and progression (54). Moreover, this class of medication is protective against cardiovascular events and has a significant role in heart failure treatment (67–69) as well as in preserving cardiac function against chemotherapy-induced cardiotoxicity (70,71). Therefore, although our analysis stratified potential confounding factors such as hypertensive diseases, heart failure, BMI, HbA1c, antihypertensive use, LEFV, NT-proBNP, and others, the observed positive association between SGLT2i and improved cancer outcomes might also reflect the enhanced control of patients overall health and comorbidities, such as those implicated in metabolic syndrome and poor cancer outcomes.

A meta-analysis of randomized controlled trials found that canagliflozin use was linked to a reduced risk of GI cancers, whereas no significant connection was observed with other SGLT2i (72). When compared to other novel anti-diabetic drugs, SGLT2i were associated with lower risks of new-onset gastric cancer (73), and colorectal cancer (74). In advanced pancreatic adenocarcinoma patients, the use of dapagliflozin concomitant with chemotherapy was well-tolerated and was suggestive of favorable changes in body composition and plasma biomarkers (75). Moreover, in a retrospective cohort study, SGLT2i use in diabetic patients with colon cancer was associated with improved survival (76). Additionally, SGLT2i initiation was suggestive of a significant reduction in liver metastatic lesions and carcinoembryonic antigen (CEA) level in a colon cancer patient, as well as spontaneous tumor regression in a patient with hepatocellular carcinoma (77,78). In light of that, our data corroborate the potential of repurposing SGLT2i for GI cancer treatment and underscore the need for prospective and randomized clinical trials that will investigate its effectiveness.

Though the contribution of our observations to the GI cancer field, this study has some limitations. Since we used retrospective EHR data, we had no control over treatment allocation, and the results reflect the treatment decisions made in the clinic. Additionally, errors in HCO reporting are possible, and some patients had missing data, such as incomplete tumor staging and ECOG status.

We also did not have information on the duration, dosage, and adherence to radiotherapy, chemotherapy, and SGLT2i treatments. On the other hand, a major strength of our study was accounting for the impact that sex differences can have on cancer outcomes and the relatively large sample of GI cancer patients who received SGLT2i while on cancer treatment.

## 5. Conclusions

In summary, SGLT2i repurposing for cancer treatment could, without major side effects, potentially improve GI cancer patients’ outcomes. However, these findings should be interpreted with caution due to the retrospective nature of the study and its inherent biases. The mechanisms underlying these effects remain unclear, and further research is needed to explore the roles of glycolytic metabolism, glycemic and insulin control, as well as the direct and indirect effects of SGLT2i on cancer (Figure 4). Additionally, new clinical trials are essential to confirm our results and explore different dosages, treatment durations, combinations, and follow-up periods, and are needed to further elucidate the potential benefits of these medications on GI cancer patients’ outcomes.

**Figure 4.**
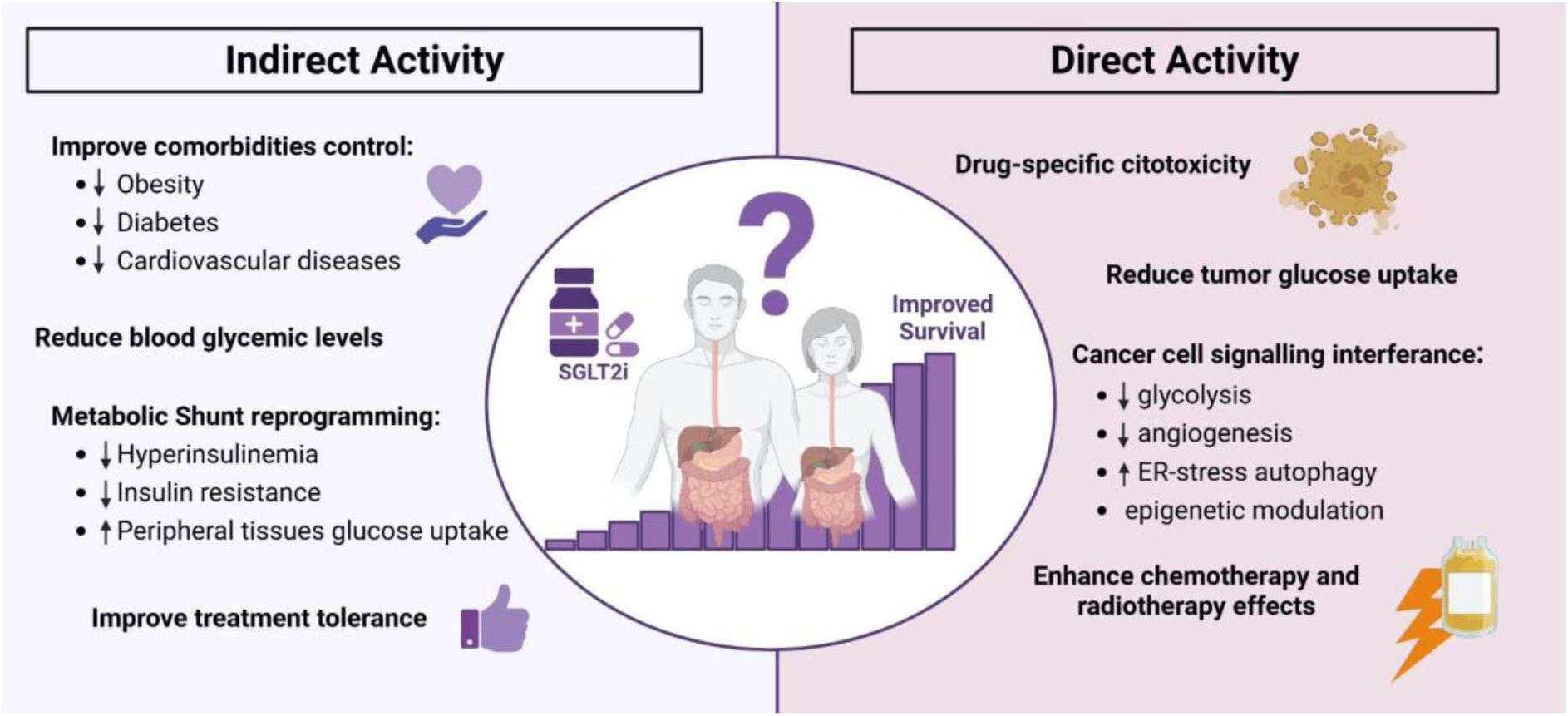
Opportunities for future research. The impact of SGLT2i on cancer survival is intriguing and warrants further investigation into their direct and indirect effects on cancer, offering promising hypotheses for future research. Although the exact mechanisms remain unclear, these inhibitors may influence tumor cells in various ways. Directly, they could exert drug-specific cytotoxic effects that promote cancer cell death. Additionally, these inhibitors might reduce glucose uptake by tumor cells, disrupt key signaling pathways involved in cell survival, proliferation, and invasion, and potentially enhance the effectiveness of chemotherapy and radiotherapy. Indirectly, they could lower tumor glucose availability and improve the overall metabolic environment by reducing glycemic levels and insulin resistance, boosting cardiovascular health, and increasing patient tolerance to cancer treatments. These potential mechanisms are interesting and further research is needed to better understand how SGLT2i might impact GI cancer survival and treatment outcomes. ER-stress: endoplasmic reticulum stress.

## Data Availability

The data analysis for this study was conducted using the built-in analytics modules of the TriNetX user portal, without direct access to the underlying data. All data was hosted by TriNetX (https://trinetx.com) and must be requested through their platform.

## 6. List of abbreviations

AKF: Acute Kidney Failure
ATC: Anatomical Therapeutic Chemical
BMI: Body-Mass Index
CEA: Carcinoembryonic Antigen
CI: Confidence Interval
CKD: Chronic Kidney Disease
CPT: Current Procedural Terminology
CVE: Cardiovascular and Cerebrovascular Events
DKA: Diabetes Ketoacidosis Events
ECOG: Eastern Cooperative Oncology Group
EHR: Electronic Health Records
ER: Endoplasmic Reticulum
GI: Gastrointestinal
GLUTs: Glucose Transporters
HR: Hazard Ratio
HCO: Healthcare Organizations
HbA1c: Hemoglobin A1C
HF: Hepatic Failure
HOSPIT: Hospitalization
HYPO: Hypoglycemic Events
IGF-I: Insulin-like Growth Factor I
IRAEs: Immune-related Adverse Events
LVEF: Left Ventricular Ejection Fraction
NT-proBNP: N-terminal Pro–B-type Natriuretic Peptide
OR: Odds Ratio
PSM: Propensity Score Matching
SGLT2: Sodium-Glucose Co-Transporter 2
SGLT2i: Sodium-Glucose Co-Transporter 2 Inhibitors
SD: Standard Deviation
ICD-10: The International Classification of Disease - Tenth Revision
TNX: TriNetX Curated Code
UTI: Urinary Tract Infections
YRS: Years

## 7. Declarations

### 7.1 Ethics approval and consent to participate

This retrospective study is exempt from informed consent. The data reviewed is a secondary analysis of existing data, does not involve intervention or interaction with human subjects, and is de-identified per the de-identification standard defined in Section §164.514(a) of the HIPAA Privacy Rule. The process by which the data is de-identified is attested to through a formal determination by a qualified expert as defined in Section §164.514(b)(1) of the HIPAA Privacy Rule. This formal determination by a qualified expert was refreshed in December 2020.

### 7.2 Consent for publication

Not applicable.

### 7.6 Authors’ contributions

LF participated in the study’s conception, wrote the original draft, collected and analyzed the data. AC and TF participated in the manuscript’s critical revision and gave input to improve its clinical relevance. WT provided access to the TriNetx platform, and participated in data analysis. RC conceived and supervised the study and participated in its critical revision. All authors read and approved the final manuscript.

## 7.7 Acknowledgements

Not applicable.

## Supplementary Material

**Table S1.** Gastrointestinal (GI) cancer types included in the analysis according to The International Classification of Disease - Tenth revision (ICD-10).

| ICD-10 - Code | Diagnosis |
| --- | --- |
| C15 | Malignant neoplasm of esophagus |
| C16 | Malignant neoplasm of stomach |
| C17 | Malignant neoplasm of small intestine |
| C18 | Malignant neoplasm of colon |
| C19 | Malignant neoplasm of rectosigmoid junction |
| C20 | Malignant neoplasm of rectum |
| C21 | Malignant neoplasm of anus and anal canal |
| C22 | Malignant neoplasm of liver and intrahepatic bile ducts |
| C23 | Malignant neoplasm of gallbladder |
| C24 | Malignant neoplasm of other and unspecified parts of biliary tract |
| C25 | Malignant neoplasm of pancreas |

**Table S2.** List of SGLT2i that patients were exposed to.

| ATC code | Name |
| --- | --- |
| A10BK01 | dapagliflozin |
| A10BK02 | canagliflozin |
| A10BK03 | empagliflozin |
| A10BK04 | ertugliflozin |
| A10BK05 | ipragliflozin |
| A10BK06 | sotagliflozin |
| A10BK07 | luseogliflozin |
| A10BK08 | bexagliflozin |

**Table S3.** Specification of the diagnoses included in the Secondary Outcomes.

| Outcomes | Codes | Definition |
| --- | --- | --- |
| Death | - | Deceased |
| Hospitalisation | CPT: 1013659 | Hospital Inpatient and Observation Care Services |
| Hypoglycemia | ICD-10-CM: E16.2 | Hypoglycemia, unspecified |
| Urinary Tract Infection | ICD-10-CM: N39.0 | Urinary tract infection, site not specified |
| Cardiovascular and cerebrovascular events | ICD-10-CM: I63 | Cerebral infarction |
|  | ICD-10-CM: I21 | Acute myocardial infarction |
| Acute Kidney Failure | ICD-10-CM: N17 | Acute kidney failure |
| Hepatic failure | ICD-10-CM: K72 | Hepatic failure, not elsewhere classified |
| Ketoacidosis | ICD-10-CM: E11.1 | Type 2 diabetes mellitus with ketoacidosis |
| Immune related adverse events | ICD-10-CM: R21 | Rash and other nonspecific skin eruption |
|  | ICD-10-CM: M06 | Other rheumatoid arthritis |
|  | ICD-10-CM: M05 | Rheumatoid arthritis with rheumatoid factor |
|  | ICD-10-CM: M35 | Other systemic involvement of connective tissue |
|  | ICD-10-CM: I77.6 | Arteritis, unspecified |
|  | ICD-10-CM: G04.9 | Encephalitis, myelitis and encephalomyelitis, unspecified |
|  | ICD-10 CM: G04.81 | Other encephalitis and encephalomyelitis |
|  | ICD-10-CM: G04.89 | Other myelitis |
|  | ICD-10-CM: E06.3 | Autoimmune thyroiditis |
|  | ICD-10-CM: K75.4 | Autoimmune hepatitis |
|  | ICD-10-CM: D59.10 | Autoimmune hemolytic anemia, unspecified |
|  | ICD-10-CM: L50.8 | Other urticaria |

CPT: current procedural terminology. ICD-10-CM: ICD-10 clinical modification.

**Table S4.** Specification of diagnoses, medications, and procedures included as covariates.

| List of diagnoses, medications, and variables included as covariates |  |
| --- | --- |
| Diagnosis |  |
| ICD-10 codes | Definition |
| E08-E13 | Diabetes mellitus |
| I10-I1A | Hypertensive diseases |
| I20-I25 | Ischemic heart diseases |
| I70-I79 | Diseases of arteries, arterioles, and capillaries |
| I60-I69 | Cerebrovascular diseases |
| I50 | Heart failure |
| N18 | Chronic kidney disease (CKD) |
| Medication |  |
| Codes | Definition |
| CV350 | antilipemic agents |
| CV100 | beta blockers/related |
| CV200 | calcium channel blockers |
| CV805 | angiotensin II inhibitor |
| CV800 | ACE inhibitors |
| CV700 | diuretics |
| HS501 | insulin |
| RxNorm 6809 | metformin |
| Laboratory |  |
| TNX: 9037 | Hemoglobin A1c/Hemoglobin.total in Blood |
| TNX: 9072 | Natriuretic peptide.B prohormone N-Terminal<br>[Mass/volume] in Serum, Plasma or Blood |
| TNX: 2003 | Left Ventricular Ejection Fraction (LVEF) (%) |
| TNX: 9083 | Body Mass Index (BMI) |
| TNX: 2002 | ECOG Performance Status |

698 TNX: TriNetX curated code; ACE: Angiotensin-Converting Enzyme.

## References

1. Arnold M, Abnet CC, Neale RE, Vignat J, Giovannucci EL, McGlynn KA, et al. Global Burden of 5 Major Types of Gastrointestinal Cancer. Gastroenterology. 2020 Jul;159(1):335–349.e15.

2. Ben-Aharon I, van Laarhoven HWM, Fontana E, Obermannova R, Nilsson M, Lordick F. Early-Onset Cancer in the Gastrointestinal Tract Is on the Rise—Evidence and Implications. Cancer Discov. 2023 Mar 1;13(3):538–51.

3. Rauth S, Malafa M, Ponnusamy MP, Batra SK. Emerging Trends in Gastrointestinal Cancer Targeted Therapies: Harnessing Tumor Microenvironment, Immune Factors, and Metabolomics Insights. Gastroenterology. 2024 Oct 1;167(5):867–84.

4. Hanahan D, Weinberg RA. Hallmarks of Cancer: The Next Generation. Cell. 2011 Mar;144(5):646–74.

5. Paul S, Ghosh S, Kumar S. Tumor glycolysis, an essential sweet tooth of tumor cells. Semin Cancer Biol. 2022 Nov 1;86:1216–30.

6. Sawayama H, Ishimoto T, Sugihara H, Miyanari N, Miyamoto Y, Baba Y, et al. Clinical impact of the Warburg effect in gastrointestinal cancer (Review). Int J Oncol. 2014 Oct 1;45(4):1345–54.

7. Icard P, Shulman S, Farhat D, Steyaert JM, Alifano M, Lincet H. How the Warburg effect supports aggressiveness and drug resistance of cancer cells? Drug Resist Updat. 2018 May 1;38:1–11.

8. Meng Y, Xu X, Luan H, Li L, Dai W, Li Z, et al. The Progress and Development of GLUT1 Inhibitors Targeting Cancer Energy Metabolism. Future Med Chem. 2019 Sep 1;11(17):2333–52.

9. Lau KTK, Ng L, Wong JWH, Loong HHF, Chan WWL, Lee CH, et al. Repurposing sodium-glucose co-transporter 2 inhibitors (SGLT2i) for cancer treatment – A Review. Rev Endocr Metab Disord. 2021 Dec 1;22(4):1121–36.

10. Koepsell H. The Na+-D-glucose cotransporters SGLT1 and SGLT2 are targets for the treatment of diabetes and cancer. Pharmacol Ther. 2017 Feb 1;170:148–65.

11. Wright EM. SGLT2 and cancer. Pflüg Arch - Eur J Physiol. 2020 Sep 1;472(9):1407– 14.

12. Scafoglio C, Hirayama BA, Kepe V, Liu J, Ghezzi C, Satyamurthy N, et al. Functional expression of sodium-glucose transporters in cancer. Proc Natl Acad Sci. 2015 Jul 28;112(30):E4111–9.

13. Perry RJ, Shulman GI. Sodium-glucose cotransporter-2 inhibitors: Understanding the mechanisms for therapeutic promise and persisting risks. J Biol Chem. 2020 Oct 16;295(42):14379–90.

14. American Diabetes Association Professional Practice Committee. 9. Pharmacologic Approaches to Glycemic Treatment: Standards of Care in Diabetes—2024. Diabetes Care. 2023 Dec 11;47(Supplement_1):S158–78.

15. Toyama T, Neuen BL, Jun M, Ohkuma T, Neal B, Jardine MJ, et al. Effect of SGLT2 inhibitors on cardiovascular, renal and safety outcomes in patients with type 2 diabetes mellitus and chronic kidney disease: A systematic review and meta-analysis. Diabetes Obes Metab. 2019 May;21(5):1237–50.

16. Patel T. SGLT2 inhibitors reduce adverse renal and CV outcomes in patients with or without diabetes. Ann Intern Med. 2023 Mar 21;176(3):JC27.

17. Rosenstock J, Aggarwal N, Polidori D, Zhao Y, Arbit D, Usiskin K, et al. Dose-Ranging Effects of Canagliflozin, a Sodium-Glucose Cotransporter 2 Inhibitor, as Add-On to Metformin in Subjects With Type 2 Diabetes. Diabetes Care. 2012 May 11;35(6):1232–8.

18. Halimi S, Vergès B. Adverse effects and safety of SGLT-2 inhibitors. Diabetes Metab. 2014 Dec 1;40(6, Supplement 1):S28–34.

19. Carlson CJ, Santamarina ML. Update review of the safety of sodium-glucose cotransporter 2 inhibitors for the treatment of patients with type 2 diabetes mellitus. Expert Opin Drug Saf. 2016 Oct 2;15(10):1401–12.

20. Scheen AJ. Pharmacodynamics, Efficacy and Safety of Sodium–Glucose Co-Transporter Type 2 (SGLT2) Inhibitors for the Treatment of Type 2 Diabetes Mellitus. Drugs. 2015 Jan 1;75(1):33–59.

21. Wiviott SD, Raz I, Bonaca MP, Mosenzon O, Kato ET, Cahn A, et al. Dapagliflozin and Cardiovascular Outcomes in Type 2 Diabetes. N Engl J Med. 2019 Jan 24;380(4):347–57.

22. Patel SM, Kang YM, Im K, Neuen BL, Anker SD, Bhatt DL, et al. Sodium-Glucose Cotransporter-2 Inhibitors and Major Adverse Cardiovascular Outcomes: A SMART-C Collaborative Meta-Analysis. Circulation. 2024 Jun 4;149(23):1789–801.

23. Brown E, Heerspink HJL, Cuthbertson DJ, Wilding JPH. SGLT2 inhibitors and GLP-1 receptor agonists: established and emerging indications. The Lancet. 2021 Jul 17;398(10296):262–76.

24. Vaduganathan M, Docherty KF, Claggett BL, Jhund PS, de Boer RA, Hernandez AF, et al. SGLT2 inhibitors in patients with heart failure: a comprehensive meta-analysis of five randomised controlled trials. The Lancet. 2022 Sep 3;400(10354):757–67.

25. Gaitonde SG, Nissan A, Protić M, Stojadinovic A, Wainberg ZA, Chen DC, et al. Sex-Specific Differences in Colon Cancer when Quality Measures Are Adhered to: Results from International, Prospective, Multicenter Clinical Trials. J Am Coll Surg. 2017 Jul;225(1):85.

26. Kalff MC, Wagner AD, Verhoeven RHA, Lemmens VEPP, van Laarhoven HWM, Gisbertz SS, et al. Sex differences in tumor characteristics, treatment, and outcomes of gastric and esophageal cancer surgery: nationwide cohort data from the Dutch Upper GI Cancer Audit. Gastric Cancer. 2022 Jan 1;25(1):22–32.

27. Baraibar I, Ros J, Saoudi N, Salvà F, García A, Castells MR, et al. Sex and gender perspectives in colorectal cancer. ESMO Open. 2023 Apr;8(2):101204.

28. Padda IS, Mahtani AU, Parmar M. Sodium-Glucose Transport Protein 2 (SGLT2) Inhibitors. In: StatPearls [Internet]. Treasure Island (FL): StatPearls Publishing; 2024 [cited 2024 Dec 1]. Available from: http://www.ncbi.nlm.nih.gov/books/NBK576405/

29. Hung MH, Chen YL, Chen LJ, Chu PY, Hsieh FS, Tsai MH, et al. Canagliflozin inhibits growth of hepatocellular carcinoma via blocking glucose-influx-induced β-catenin activation. Cell Death Dis. 2019 May 29;10(6):1–15.

30. Kaji K, Nishimura N, Seki K, Sato S, Saikawa S, Nakanishi K, et al. Sodium glucose cotransporter 2 inhibitor canagliflozin attenuates liver cancer cell growth and angiogenic activity by inhibiting glucose uptake. Int J Cancer. 2018;142(8):1712–22.

31. Nakano D, Kawaguchi T, Iwamoto H, Hayakawa M, Koga H, Torimura T. Effects of canagliflozin on growth and metabolic reprograming in hepatocellular carcinoma cells: Multi-omics analysis of metabolomics and absolute quantification proteomics (iMPAQT). PLOS ONE. 2020 Apr 28;15(4):e0232283.

32. Luo J, Sun P, Zhang X, Lin G, Xin Q, Niu Y, et al. Canagliflozin Modulates Hypoxia - Induced Metastasis, Angiogenesis and Glycolysis by Decreasing HIF-1α Protein Synthesis via AKT/mTOR Pathway. Int J Mol Sci. 2021 Jan;22(24):13336.

33. Abdel-Rafei MK, Thabet NM, Rashed LA, Moustafa EM. Canagliflozin, a SGLT-2 inhibitor, relieves ER stress, modulates autophagy and induces apoptosis in irradiated HepG2 cells: Signal transduction between PI3K/AKT/GSK-3β/mTOR and Wnt/β-catenin pathways;: in vitro. J Cancer Res Ther. 2021 Dec;17(6):1404.

34. Zeng Y, Jiang H, Zhang X, Xu J, Wu X, Xu Q, et al. Canagliflozin reduces chemoresistance in hepatocellular carcinoma through PKM2-c-Myc complex-mediated glutamine starvation. Free Radic Biol Med. 2023 Nov 1;208:571–86.

35. Zhong J, Sun P, Xu N, Liao M, Xu C, Ding Y, et al. Canagliflozin inhibits p-gp function and early autophagy and improves the sensitivity to the antitumor effect of doxorubicin. Biochem Pharmacol. 2020 May 1;175:113856.

36. Zhou J, Feng J, Wu Y, Dai HQ, Zhu GZ, Chen PH, et al. Simultaneous treatment with sorafenib and glucose restriction inhibits hepatocellular carcinoma in vitro and in vivo by impairing SIAH1-mediated mitophagy. Exp Mol Med. 2022 Nov;54(11):2007–21.

37. Xu D, Zhou Y, Xie X, He L, Ding J, Pang S, et al. Inhibitory effects of canagliflozin on pancreatic cancer are mediated via the downregulation of glucose transporter-1 and lactate dehydrogenase A. Int J Oncol. 2020 Nov 1;57(5):1223–33.

38. Ren D, Sun Y, Zhang D, Li D, Liu Z, Jin X, et al. SGLT2 promotes pancreatic cancer progression by activating the Hippo signaling pathway via the hnRNPK-YAP1 axis. Cancer Lett. 2021 Oct 28;519:277–88.

39. Nasiri AR, Rodrigues MR, Li Z, Leitner BP, Perry RJ. SGLT2 inhibition slows tumor growth in mice by reversing hyperinsulinemia. Cancer Metab. 2019 Dec 11;7(1):10.

40. Okada J, Yamada E, Saito T, Yokoo H, Osaki A, Shimoda Y, et al. Dapagliflozin Inhibits Cell Adhesion to Collagen I and IV and Increases Ectodomain Proteolytic Cleavage of DDR1 by Increasing ADAM10 Activity. Molecules. 2020 Jan;25(3):495.

41. Anastasio C, Donisi I, Del Vecchio V, Colloca A, Mele L, Sardu C, et al. SGLT2 inhibitor promotes mitochondrial dysfunction and ER-phagy in colorectal cancer cells. Cell Mol Biol Lett. 2024 May 29;29(1):80.

42. Ren K, Wang X, Ma R, Chen H, Min T, Ma Y, et al. Dapagliflozin suppressed gastric cancer growth via regulating OTUD5 mediated YAP1 deubiquitination. Eur J Pharmacol. 2024 Nov 15;983:177002.

43. Jiang D, Ma P. Canagliflozin, characterized as a HDAC6 inhibitor, inhibits gastric cancer metastasis. Front Oncol. 2022 Nov 15;12:1057455.

44. Roderburg C, Loosen SH, Hoyer L, Luedde T, Kostev K. Prevalence of diabetes mellitus among 80,193 gastrointestinal cancer patients in five European and three Asian countries. J Cancer Res Clin Oncol. 2022 May 1;148(5):1057–62.

45. Yang K, Liu Z, Thong MSY, Doege D, Arndt V. Higher Incidence of Diabetes in Cancer Patients Compared to Cancer-Free Population Controls: A Systematic Review and Meta-Analysis. Cancers. 2022 Jan;14(7):1808.

46. Lawler T, Walts ZL, Steinwandel M, Lipworth L, Murff HJ, Zheng W, et al. Type 2 Diabetes and Colorectal Cancer Risk. JAMA Netw Open. 2023 Nov 14;6(11):e2343333.

47. Yoon JM, Son KY, Eom CS, Durrance D, Park SM. Pre-existing diabetes mellitus increases the risk of gastric cancer: A meta-analysis. World J Gastroenterol. 2013 Feb 14;19(6):936–45.

48. Cheung KS, Chan EW, Chen L, Seto WK, Wong ICK, Leung WK. Diabetes Increases Risk of Gastric Cancer After Helicobacter pylori Eradication: A Territory-Wide Study With Propensity Score Analysis. Diabetes Care. 2019 Jul 11;42(9):1769–75.

49. Sekikawa A, Fukui H, Maruo T, Tsumura T, Okabe Y, Osaki Y. Diabetes mellitus increases the risk of early gastric cancer development. Eur J Cancer. 2014 Aug 1;50(12):2065–71.

50. Andersen DK, Korc M, Petersen GM, Eibl G, Li D, Rickels MR, et al. Diabetes, Pancreatogenic Diabetes, and Pancreatic Cancer. Diabetes. 2017 May 1;66(5):1103–10.

51. Conway RBN, Sudenga S, McClain D, Blot WJ. Diabetes and liver cancer risk: A stronger effect in Whites than Blacks? J Diabetes Complications. 2021 Mar 1;35(3):107816.

52. Lee SJ, Kim C, Yu H, Kim DK. Analysis of the Incidence of Type 2 Diabetes, Requirement of Insulin Treatment, and Diabetes-Related Complications among Patients with Cancer. Cancers. 2023 Jan;15(4):1094.

53. Sylow L, Grand MK, von Heymann A, Persson F, Siersma V, Kriegbaum M, et al. Incidence of New-Onset Type 2 Diabetes After Cancer: A Danish Cohort Study. Diabetes Care. 2022 May 27;45(6):e105–6.

54. Gallagher EJ, LeRoith D. Obesity and Diabetes: The Increased Risk of Cancer and Cancer-Related Mortality. Physiol Rev. 2015 Jul;95(3):727–48.

55. Campbell PT, Newton CC, Patel AV, Jacobs EJ, Gapstur SM. Diabetes and Cause-Specific Mortality in a Prospective Cohort of One Million U.S. Adults. Diabetes Care. 2012 Aug 14;35(9):1835–44.

56. Han X, Raun SH, Carlsson M, Sjøberg KA, Henriquez-Olguín C, Ali M, et al. Cancer causes metabolic perturbations associated with reduced insulin-stimulated glucose uptake in peripheral tissues and impaired muscle microvascular perfusion. Metab - Clin Exp [Internet]. 2020 Apr 1 [cited 2024 Dec 24];105. Available from: https://www.metabolismjournal.com/article/S0026-0495(20)30033-0/abstract

57. Masi T, Patel BM. Altered glucose metabolism and insulin resistance in cancer-induced cachexia: a sweet poison. Pharmacol Rep. 2021 Feb 1;73(1):17–30.

58. Asp ML, Tian M, Wendel AA, Belury MA. Evidence for the contribution of insulin resistance to the development of cachexia in tumor-bearing mice. Int J Cancer. 2010;126(3):756–63.

59. Ferrer M, Anthony TG, Ayres JS, Biffi G, Brown JC, Caan BJ, et al. Cachexia: A systemic consequence of progressive, unresolved disease. Cell. 2023 Apr 27;186(9):1824– 45.

60. Gilmore LA, Olaechea S, Gilmore BW, Gannavarapu BS, Alvarez CM, Ahn C, et al. A preponderance of gastrointestinal cancer patients transition into cachexia syndrome. J Cachexia Sarcopenia Muscle. 2022;13(6):2920–31.

61. Martin L, Birdsell L, MacDonald N, Reiman T, Clandinin MT, McCargar LJ, et al. Cancer Cachexia in the Age of Obesity: Skeletal Muscle Depletion Is a Powerful Prognostic Factor, Independent of Body Mass Index. J Clin Oncol. 2013 Apr 20;31(12):1539–47.

62. Erdiş E. Does Diabetes Mellitus Increase Radiotherapy/ Chemoradiotherapy Acute Toxicities? Turk J Oncol. 2023;466–75.

63. Mailliez A, Ternynck C, Duhamel A, Mailliez A, Ploquin A, Desauw C, et al. Diabetes is associated with high risk of severe adverse events during chemotherapy for cancer patients: A single-center study. Int J Cancer. 2022 Sep 14;152(3):408.

64. Lee PC, Ganguly S, Goh S -Y. Weight loss associated with sodium-glucose cotransporter-2 inhibition: a review of evidence and underlying mechanisms. Obes Rev. 2018 Dec;19(12):1630–41.

65. Teo YH, Teo YN, Syn NL, Kow CS, Yoong CSY, Tan BYQ, et al. Effects of Sodium/Glucose Cotransporter 2 (SGLT2) Inhibitors on Cardiovascular and Metabolic Outcomes in Patients Without Diabetes Mellitus: A Systematic Review and Meta-Analysis of Randomized-Controlled Trials. J Am Heart Assoc. 2021 Mar 2;10(5):e019463.

66. Georgianos PI, Agarwal R. Ambulatory Blood Pressure Reduction With SGLT-2 Inhibitors: Dose-Response Meta-analysis and Comparative Evaluation With Low-Dose Hydrochlorothiazide. Diabetes Care. 2019 Mar 11;42(4):693–700.

67. Zannad F, Ferreira JP, Pocock SJ, Anker SD, Butler J, Filippatos G, et al. SGLT2 inhibitors in patients with heart failure with reduced ejection fraction: a meta-analysis of the EMPEROR-Reduced and DAPA-HF trials. The Lancet. 2020 Sep 19;396(10254):819–29.

68. Usman MS, Siddiqi TJ, Anker SD, Bakris GL, Bhatt DL, Filippatos G, et al. Effect of SGLT2 Inhibitors on Cardiovascular Outcomes Across Various Patient Populations. J Am Coll Cardiol. 2023 Jun 27;81(25):2377–87.

69. Usman MS, Bhatt DL, Hameed I, Anker SD, Cheng AYY, Hernandez AF, et al. Effect of SGLT2 inhibitors on heart failure outcomes and cardiovascular death across the cardiometabolic disease spectrum: a systematic review and meta-analysis. Lancet Diabetes Endocrinol. 2024 Jul 1;12(7):447–61.

70. Dabour MS, George MY, Daniel MR, Blaes AH, Zordoky BN. The Cardioprotective and Anticancer Effects of SGLT2 Inhibitors. JACC CardioOncology. 2024 Apr;6(2):159–82.

71. Gongora CA, Drobni ZD, Quinaglia Araujo Costa Silva T, Zafar A, Gong J, Zlotoff DA, et al. Sodium-Glucose Co-Transporter-2 Inhibitors and Cardiac Outcomes Among Patients Treated With Anthracyclines. JACC Heart Fail. 2022 Aug 1;10(8):559–67.

72. Tang H, Dai Q, Shi W, Zhai S, Song Y, Han J. SGLT2 inhibitors and risk of cancer in type 2 diabetes: a systematic review and meta-analysis of randomised controlled trials. Diabetologia. 2017 Oct 1;60(10):1862–72.

73. Chou OHI, Chauhan VK, Chung CTS, Lu L, Lee TTL, Ng ZMW, et al. Comparative effectiveness of sodium-glucose cotransporter-2 inhibitors for new-onset gastric cancer and gastric diseases in patients with type 2 diabetes mellitus: a population-based cohort study. Gastric Cancer. 2024 Sep 1;27(5):947–70.

74. Suzuki Y, Kaneko H, Okada A, Ko T, Jimba T, Fujiu K, et al. Association of SGLT2 inhibitors with incident cancer. Diabetes Metab. 2024 Nov 1;50(6):101585.

75. Park LK, Lim KH, Volkman J, Abdiannia M, Johnston H, Nigogosyan Z, et al. Safety, tolerability, and effectiveness of the sodium-glucose cotransporter 2 inhibitor (SGLT2i) dapagliflozin in combination with standard chemotherapy for patients with advanced, inoperable pancreatic adenocarcinoma: a phase 1b observational study. Cancer Metab. 2023 May 18;11(1):6.

76. Chiang CH, Chiang CH, Hsia YP, Jaroenlapnopparat A, Horng CS, Wong KY, et al. The impact of sodium-glucose cotransporter-2 inhibitors on outcome of patients with diabetes mellitus and colorectal cancer. J Gastroenterol Hepatol. 2024;39(5):902–7.

77. Okada J, Matsumoto S, Kaira K, Saito T, Yamada E, Yokoo H, et al. Sodium Glucose Cotransporter 2 Inhibition Combined With Cetuximab Significantly Reduced Tumor Size and Carcinoembryonic Antigen Level in Colon Cancer Metastatic to Liver. Clin Colorectal Cancer. 2018 Mar 1;17(1):e45–8.

78. Kawaguchi T, Nakano D, Okamura S, Shimose S, Hayakawa M, Niizeki T, et al. Spontaneous regression of hepatocellular carcinoma with reduction in angiogenesis-related cytokines after treatment with sodium-glucose cotransporter 2 inhibitor in a cirrhotic patient with diabetes mellitus. Hepatol Res. 2019;49(4):479–86.

